# A Randomised Controlled Trial of Nasal Immunisation with Live Virulence Attenuated *Streptococcus pneumoniae* Strains using Human Infection Challenge

**DOI:** 10.1101/2023.04.14.23288224

**Authors:** H Hill, E Mitsi, E Nikolaou, A Blizard, S Pojar, A Howard, A Hyder-Wright, Jack Devin, J Reiné, R Robinson, C Solórzano, S Jochems, T Kenny-Nyazika, E Ramos-Sevillano, CM Weight, C Myerscough, D McLennan, B Morton, E Gibbons, M Farrar, V Randles, H Burhan, T Chen, AD Shandling, JJ Campo, R Heyderman, SB Gordon, J Brown, AM Collins, DM Ferreira

**Author notes:** Authors contriduted equally to this work (H Hill, E Mitsi). joint senior authors (J Brown, AM Collins, DM Ferreira). **ISRCTN registry:** ISRCTN22467293. **Author contributions** -Study concept and design: HH, EM, RH, SBG, JB, AMC, DMF. -Study protocol writing, ethics submission and study co-ordination: HH, AHW, EM, EN, JB, SBG, AMC, DMF -Clinical cover including on call responsibility: RR, DMcL, BM, EG, VR, HB, AMC - Recruiting and consenting participants: AHW, RR, DMcL, DC, EG, MF, VR - Provision of reagents, bacteria and expertise: ERS, CMW, JB, RH, JJC - Bacterial inoculum preparation, sample processing and data collection: HH, EM, AB, EN, SP, AH, JR, RR, CS, SJ, TKN, ERS, CMW, DL, CM, DMcL, DC, EG, MF, VR, ADS, JJC - Data analysis and interpretation: HH, EM, TKN, JJC, RH, JB, AMC, DMF - Statistical planning and analysis: HH, EM, CT, DMF - manuscript preparation and review: all authors. **Funding:** Funded by Bill and Melinda Gates Foundation (OPP1117728) and the Medical Research Council awards (MR/M011569/1 and MR/N016874/1). JSB, RH, CMW and ERS work at UCLH/UCL which received funding from the Department of Health’s NIHR Biomedical Research Centre’s funding scheme. RSH is a NIHR Senior Investigator.

## Abstract

**Rationale:** Pneumococcal pneumonia remains a global health problem. Pneumococcal colonisation increases local and systemic protective immunity, suggesting nasal administration of live attenuated *S. pneumoniae* strains could help prevent infections.

**Objectives:** We used a controlled human infection model to investigate whether nasopharyngeal colonisation with attenuated *S. pneumoniae* strains protected against re-colonisation with wild-type (WT) *S. pneumoniae* (Spn).

**Methods:** Healthy adults aged 18-50 years were randomised (1:1:1:1) for nasal administration twice (two weeks interval) with saline, WT Spn6B (BHN418) or one of two genetically modified Spn6B strains - SpnA1 (Δ*fhs/piaA*) or SpnA3 (Δ*proABC/piaA*) (Stage I). After 6 months, participants were challenged with SpnWT to assess protection against re-colonisation (Stage II).

**Measurements and Main Results:** 125 participants completed both study stages as per intention to treat. No Serious Adverse Events were reported. In Stage I, colonisation rates were similar amongst groups: SpnWT 58.1% (18/31), SpnA1 60% (18/30) and SpnA3 59.4% (19/32). Anti-Spn nasal IgG levels post-colonisation were similar in all groups whilst serum IgG responses were higher in the SpnWT and SpnA1 groups than the SpnA3 group. In colonised individuals, increases in IgG responses were identified against 197 Spn protein antigens and serotype 6 capsular polysaccharide using a pangenome array. Participants given SpnWT or SpnA1 but not SpnA3 in phase 1 were partially protected against re-colonisation with SpnWT (recolonisation rates of 29% versus 30% respectively).

**Conclusion:** Nasal colonisation with genetically modified live attenuated Spn was safe and induced protection against recolonisation, suggesting nasal adminstration of live attenuated Spn could be an effective stategy for preventing pneumococcal infections.

## Introduction

*Streptococcus pneumoniae* (Spn) is the dominant bacterial pathogen causing acute lower respiratory tract (LRTI) infections in adults, responsible for up to 50% of community acquired pneumonia (1-3) and 25% of COPD exacerbations (4). Despite this major clinical need for preventing pneumococcal infections, existing vaccines have significant drawbacks. The pneumococcal polysaccharide vaccine (PPV) used in adults has limited efficacy at preventing Spn lung infections (5). Although the conjugated polysaccharide vaccine (PCV) used routinely in children is effective against pneumonia, it only protects against a limited number of capsular serotypes and its high price limit PCV widead use in low- and middle-income countries without GAVI support (6, 7). Furthermore, a high proportion of adult disease is caused by non-vaccine serotypes (8), the prevalence of which is increasing due to serotype replacement in response to infant vaccination (8, 9).

Extensive human and mouse data demonstrate that Spn nasopharyngeal colonisation is an immunising event, inducing antibodies to capsular antigens and antibody and T cell responses to protein antigens in the periphery and local mucosa. These immune responses can prevent re-colonisation of the nasopharynx and thereby prevent development of invasive disease (10-24). As a consequence, all adults have developed naturally acquired immunity against Spn that can be boosted by natural re-colonisation events (19, 22, 23). These data suggest a novel preventative strategy could be the deliberate nasopharyngeal administration of live Spn aiming to boost existing naturally acquired immunity, which could prevent re-colonisation with virulent Spn and enhance protective immunity against pneumonia and systemic infection. A similar strategy has been proposed for prevention of *Neisseria meningitidis* meningitis by nasal administration of the avirulent related species Neisseria lactamica (25, 26). Our previous pre-clinical data demonstrated that the double mutant Spn serotype 6B *ΔproABC/piaA* and *Δfhs/piaA* strains are suitable to explore this strategy. In murine infection models both *ΔproABC/piaA* and *Δfhs/piaA* were markedly attenuated in virulence and therefore unlikely to cause disease in a target population with increased susceptibility to Spn. Despite this, in mouse models nasopharyngeal administration with either strain stimulated significant protective adaptive immunity against subsequent colonisation, pneumonia and sepsis with the homologous wild-type serotype 6B (24).

Using the well established Experimental Human Pneumococcal Challenge (EHPC) model (22, 27-30) we have previously described the human immunological responses to nasopharyngeal colonisation with Spn, and demonstrated the efficacy of PCV-13 vaccination in preventing colonisation (22, 31, 32). Here, we have used the EHPC model to test whether nasopharyngeal administration of the Spn6B Δfhs/piaA or Spn6B ΔproABC/piaA strain prevent subsequent recolonisation with wild-type Spn. As secondary endpoints, we assessed the induction of antibody responses against protein and capsular polysaccharide Spn antigens following experimental colonisation.

## Methods

### Trial Design and Participants

The study was a single-blind randomised controlled clinical trial with an adaptive design using the Experimental Human Pneumococcal Colonisation (EHPC) model. The study was conducted in 2018-2019 at the Liverpool School of Tropical Medicine, Liverpool, UK. Participants were healthy adults aged 18 to 50 years (n=193), consented and then screened to exclude health conditions that confound the study outcome including risk factors for infection for participants, their close contacts or pregnancy (Supplementary Figure 1). In Stage I, participants were randomised to one of three study arms for nasal inoculation with 80,000 CFU per nostril on day 0 followed by a second dose on day 14 (Figure 1A) whereas the fourth arm received inoculation with saline twice. Intervention arms included the wild type Spn6B BHN418 strain (SpnWT)(22), an Spn double mutant strain constructed on the backbone of BHN418 strain [SpnA1 (*Δfhs/piaA*) or initially SpnA2 (Δ*cps/proABC*)] or saline. SpnA2 was subsequently replaced due to futility (failure to colonise) by SpnA3 (ΔproABC/piaA). All participants were prescribed Amoxicillin 500mg for 3 days at the end of Stage I. In Stage II, participants were re-screened and then challenged with 80,000 CFU per nostril of wild type Spn6B at 22 weeks (min-max range: 12 and 52 weeks, respectively) after the first inoculation. Nasal wash (NW) and blood samples were obtained at follow-up visits to assess Spn nasopharyngeal colonisation and immune responses. Naturally acquired Spn colonisation in Stage I was not an exclusion criterium. Participants with persisting Spn colonisation at the end of Stage I were excluded from analysis (Supplementary Table 1). An Independent Data Monitoring and Safety Committee (DMSC) provided trial oversight and authorised the trial continuation after interim analysis of colonisation and safety data for the first 10 participants.

**Figure 1.**
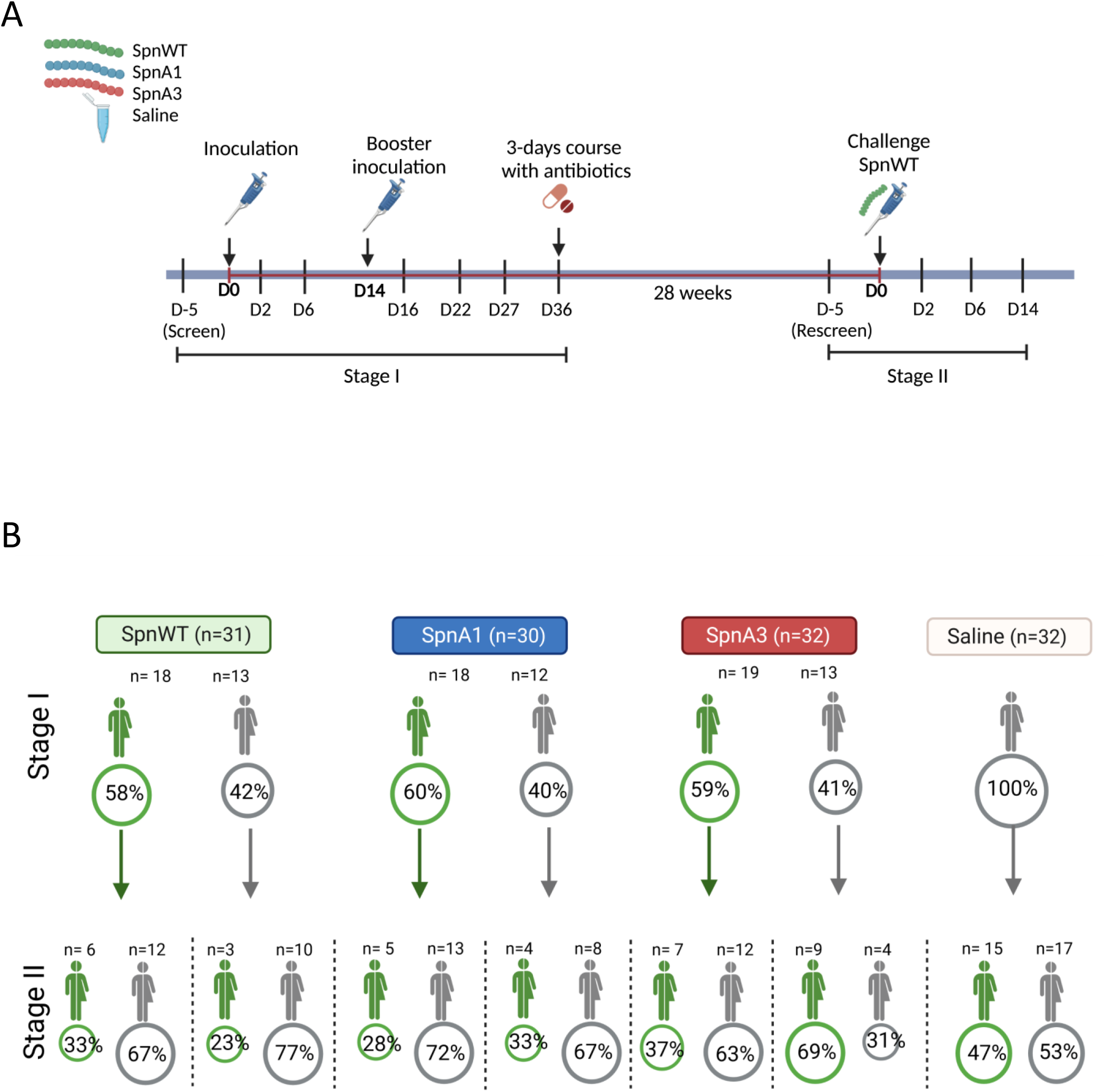
**(A) Study design**. Stage I: Pneumococcal inoculation with SpnWT or an attenuated strain (D0) and booster inoculation (14 days later). Stage II: Challenge with SpnWT at 22 weeks post initial inoculation episode (irrespective of colonisation status during Stage I). **(B) Colonisation rates**. Numbers (n) and percentages of participants colonised in each group in Stage I and Stage II.

### Regulatory and Ethical Approvals

Approvals were given by the Health Research Authority National Research Ethics Service Liverpool East (18/NW/0481) and the Department for Food Rural and Agricultural Affairs (DEFRA) for the deliberate release of a GMO under schedule 2 of the Genetically Modified (Deliberate Release) Regulations 2002 Ref 18/R51/01.

### Randomisation and Blinding

Permuted block randomisation method was implemented using SAS PROC PLAN. Computer-generated blocks of 5 with blinded envelopes were produced by an independent statistician at LSTM. After completion of Stage I, the clinical team were unblinded, then the participants unique, non-identifiable study number was changed for Stage II to ensure the laboratory scientists were blind to the initial allocation.

### Participant Monitoring and Safety

Established EHPC safety guidelines include screening to ensure volunteers are healthy then training participants to identify and respond to symptoms early supported by a safety leaflet, 24/7 clinical on call service and adverse events records as per protocol. Temperature and symptoms were reported systematically for 3 days post-inoculation and at follow up visits. A precaution for 3-days course of Amoxicillin 500mg was provided to participants on the day of inoculation to use as treatment if they developed symptoms, ideally after discussion with the on-call clinician.

### Experimental Human Pneumococcal Inoculation and Detection of Colonisation

Pneumococcal stocks for inoculation were grown to early-to-mid-log phase in vegetone as previously described (32, 33). Serotyping, penicillin and other antibiotic sensitivity were independently confirmed by Public Health England (PHE). Pre-prepared stocks were diluted on the day of inoculation to a dose of 80,000 CFU/100 μl per nostril. A multiplex PCR targeting lytA, the antibiotic resistant gene, or the deleted genes confirmed the mutant strain identities. NW samples were collected at days -5 (screen), 2, 6, 16, 22, 27 and 36 post first inoculation dose (Stage I) and at days -5 (re-screen), 2, 7 and 14 post challenge (Stage II) (Figure 1A), and plated on blood agar plates supplemented with either gentamycin or selective antibiotic for mutant strains (spectinomycin/kanamycin) and overnight incubation at 37°C in 5% CO2. Colonies were confirmed as Spn by classical methods including: (i) typical draughtsman-like colony morphology, (ii) the presence of α-haemolysis, (iii) optochin sensitivity and (iv) solubility in bile salts, and serotyped using a latex agglutination kit (Staten Serum Institute).

### Pneumococcal Whole Cell ELISA

Antibody titres to whole cell Spn6B strain were determined in serum and nasal wash samples as previously described (14). Briefly, 100μl of 10^8^ CFU/ml bacterial cells prepared in carbonate buffer (pH 8) were added to the 96-well plates (Maxipore, Nunc) and allowed to adhere for 16 hours at 4 C. Plates were washed 3 times with PBS plus 0.005% Tween 20 (MilliporeSigma), and then samples were incubated for 2 hours at room temperature. All samples were run in duplicates in three 1/3 serial dilutions, starting from 1:2000 for serum and 1:2 for NW samples. Serum and nasal IgG to Spn antigens were detected using alkaline-phosphatase conjugated anti-human IgG (Sigma, A9544) and p-nitrophenyl phosphate (PNPP) as the substrate. Optical density (OD) was measured at 405 nm using a FLUOstar Omega plate reader (BMG Labtech, UK), the average blank corrected value was calculated for each sample, and data analysed using Omega Analysis (BMG Labtech).

### Serum anti-pneumococcal capsular polysaccharide antibodies

Serum IgG to Spn6B capsular polysaccharides was measured using the modified WHO standardized enzyme-linked immunosorbent assay (ELISA), as previously described (22, 28). Serum samples were run in duplicate 1:100 dilutions followed by two 1/3 serial dilutions. Antigen specific antibodies (anti-CPS6B IgG) were detected using goat anti-human alkaline phosphatase (Sigma, A9544) and p-nitrophenyl phosphate (PNPP). OD was measured at 405 nm using FLUOstar Omega plate reader (BMG Labtech).

### Pneumococcal proteome-wide and capsular polysaccharide antibodies

The Spn pangenome microarray (Antigen Discovery, Inc. [ADI], Irvine, CA, USA), included 2,036 proteins identified in clinical isolates from the Massachusetts SPARC study (34), 301 non-redundant TIGR4 reference strain proteins, 117 “diverse core loci” (including 34 pspA, 47 pspC, 19 zmpA and 17 zmpB alleles) (35), and 24 purified capsular polysaccharides (serotypes 1, 2, 3, 4, 5, 6A, 6B, 7F, 8, 9N, 9V, 10A, 11A, 12F, 14, 15B, 17F, 18C, 19A, 19F, 20, 22F, 23F and 33F) totalling 2,677 spots, including overlapping fragments for proteins longer than 1,000 amino acids, in two dilutions (0.03 µg/ml and 0.1 µg/ml). Pneumococcal proteins were expressed using an E. coli in vitro transcription and translation (IVTT) system (Rapid Translation System, Biotechrabbit, Berlin, Germany) and printed onto nitrocellulose-coated glass AVID slides (Grace Bio-Labs, Inc., Bend, OR, USA) using an Omni Grid Accent robotic microarray printer (Digilabs, Inc., Marlborough, MA, USA). Microarrays were probed with sera and antibody binding detected by incubation with DyLight650 fluorochrome-conjugated goat anti-human IgG (Bethyl Laboratories, Inc., Montgomery, TX, USA, Cat# A80-104D5). Slides were scanned on a GenePix 4300A High-Resolution Microarray Scanner (Molecular Devices, Sunnyvale, CA, USA), and raw spot and local background fluorescence intensities, spot annotations and sample phenotypes were imported and merged in R statistical software, in which all subsequent procedures were performed. Foreground spot intensities were adjusted by subtraction of local background, and negative values were converted to one. All foreground values were transformed using the base two logarithm. The dataset was normalized to remove systematic effects by subtracting the median signal intensity of the IVTT controls for each sample i.e. a value of 0.0 means the intensity is no different than background, and a value of 1.0 indicates double the background intensity.

### Statistical Methods and Analysis

The primary endpoint was protection against SpnWT colonisation at Stage II, defined as the positive culture of SpnWT in nasal wash at any time point after rechallenge. Secondary endpoints assessed density and duration of colonisation, and immunological responses. Power calculations indicated 27 participants per group were required to detect a reduction in the proportion of participants colonised after SpnWT challenge (Stage II) from 50% (negative control group) to 15% for the active groups (80% power at two sided 5% alpha level). Modified intention to treat (mITT) population was defined as participants who were randomised in Stage I and then challenged in Stage II with SpnWT. Participants who remained experimentally colonised with 6B strain when re-screened for Stage II (N=2 participants) were excluded. The primary outcome was summarised as number (%) of colonised participants and the risk ratio with 95% confidence intervals (CI), and analysed using the generalized linear model with binomial distribution and log link function based on the mITT population. For pneumococcal colonisation density at different time points, log transformed values [log10(density +1)] were analysed using GEE models with a normal distribution and identity link function. The geometric mean ratio with 95% CI between active group and placebo group was derived. Missing densities were interpolated from the mean of flanking density values or if sequential then extrapolated using the average change over that interval. Analyses were performed using SAS vs. 9.4 (SAS Institute Inc, Cary, NC). Longitudinal antibody data were analysed using ANOVA test. Protein microarray data were analysed using paired t-tests for each antigen on the array. Adjustment for the false discovery rate was performed using the “p.adjust” function in R using the method described by Benjamini and Hochberg (36). Data visualisation was performed using the ggplot2 package in R.

## Results

### Trial participants and inoculation doses

148 participants were randomised and inoculated twice in Stage I with either SpnWT (n=35), saline (n=34), SpnA1 (n=35), SpnA2 (n=9) and SpnA3 (n=35) (Supplementary Figure 1). The interim analysis reported A2 (n=9) had very low rates of colonisation (1/9, 11%) which failed to result in increased antibody levels (Supplementary Figure 2); therefore A2 was replaced by A3. After approximately 150 days, participants were challenged with SpnWT (Stage II). Data on re-colonisation were analysed from 125 participants according to the MITT (SpnWT n=31, SpnA1 n=30, SpnA3 n=32, Saline n=32) (Supplementary Figure 1). Fourteen participants were excluded from analysis as they were lost at follow up, used antibiotics during follow up, or were persistingly colonised with Spn at the time of inoculation in Stage II. Demographics were similar for all groups (Supplementary Table 2), with a median age in years of the SpnWT group of 22 (IQR:20-23), SpnA1 group of 21 (IQR:19-2), SpnA3 group of 21.5 (IQR:19-24.5), and saline group of 22 (IQR:20.5-25). The proportion of females were: SpnWT 68%, SpnA1 50%, SpnA3 72%, and saline 66%. Median doses for first and booster inoculations and Stage II WT challenge were similar between all active groups. The interval between first inoculation and booster was 14 days, and to Stage II WT rechallenge was 154.0 (IQR: 146.7-166.4) days with no differences between the groups (Supplementary Table 2).

### Similar rates and duration of colonisation by the SpnWT and mutant strains SpnA1 and SpnA3

In Stage I, overall colonisation rates following double inoculation with the SpnWT or SpnA1 or SpnA3 strains were similar amongst groups, with colonisation (at any time point) detected in 58.1% (18/31) for WT, 60% (18/30) for SpnA1 and 59.4% (19/32) for A3 (Table 1, Figure 1B). The duration of colonisation was also similar with a median of 36 days of colonisation (IQR 22-36) for all three active groups (Supplementary Table 3). However, the density of colonisation showed some differences. At day 2 the SpnWT group had increased colonisation density compared to SpnA3 (WT: 2.45 ± 1.64 log10 CFU/ml NW vs A3:1.08 ± 0.78 log10 CFU/ml NW) (p=0.03 by one-way Anova test) (Figure 2A and Supplementary Table 4). Also, the area under the curve (AUC) of colonisation density for the days 2 to day 6 interval was significantly higher for the SpnWT group compared to SpnA3 (WT: 7.13 ± 5.39 vs A3: 3.91 ± 3.02, p <0.05 by one-way Anova test) and for the SpnA1 group compared to A3 for the day 2 to day 16 interval (A1: 22.05 ± 16.77 vs A3: 10.59 ± 10.78, p <0.05, by one-way Anova Test)(Figure 2B, Supplementary Table 5).

**Table 1:**
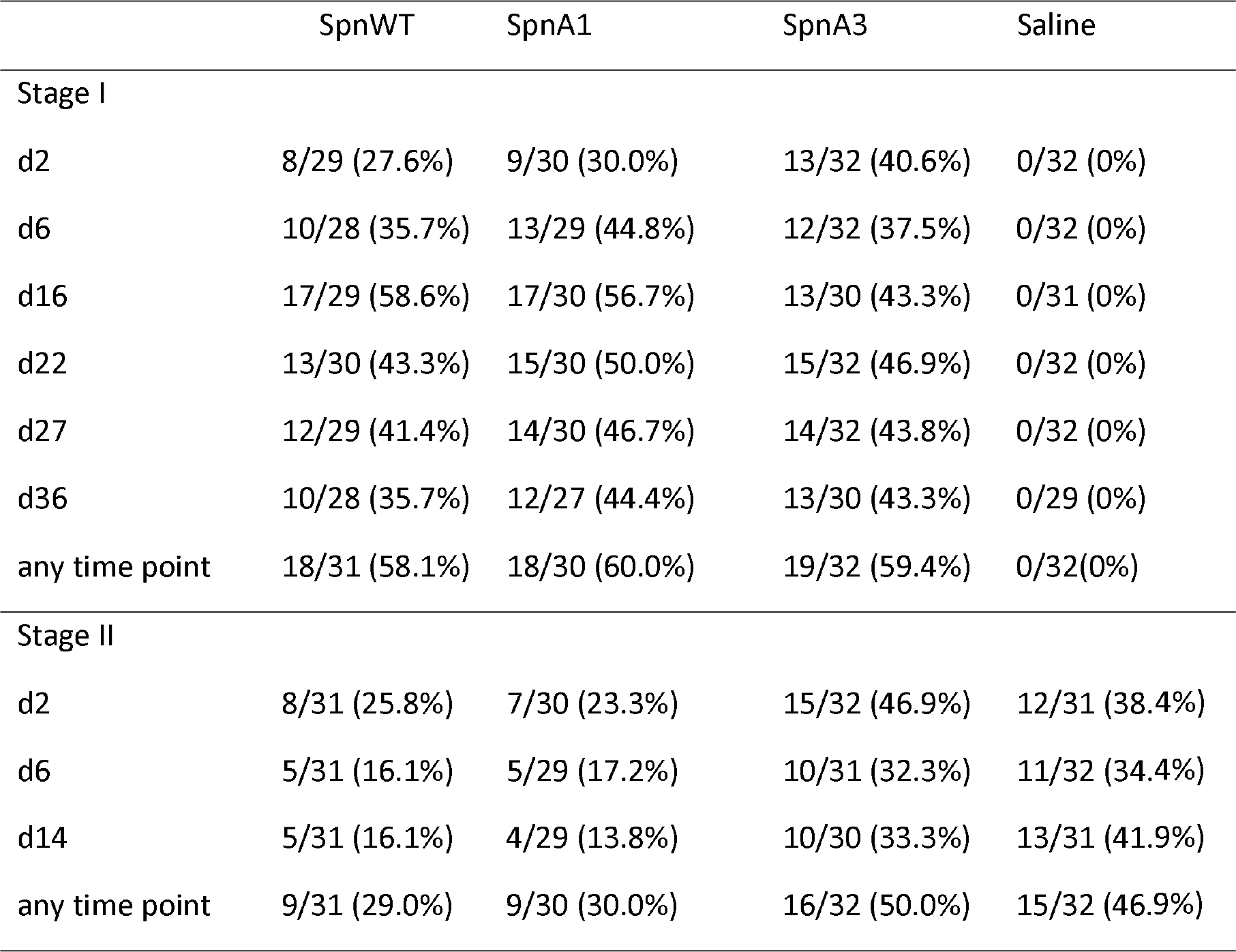
Number of participants colonised at each timepoint (defined as having a positive nasal wash culture)

**Figure 2.**
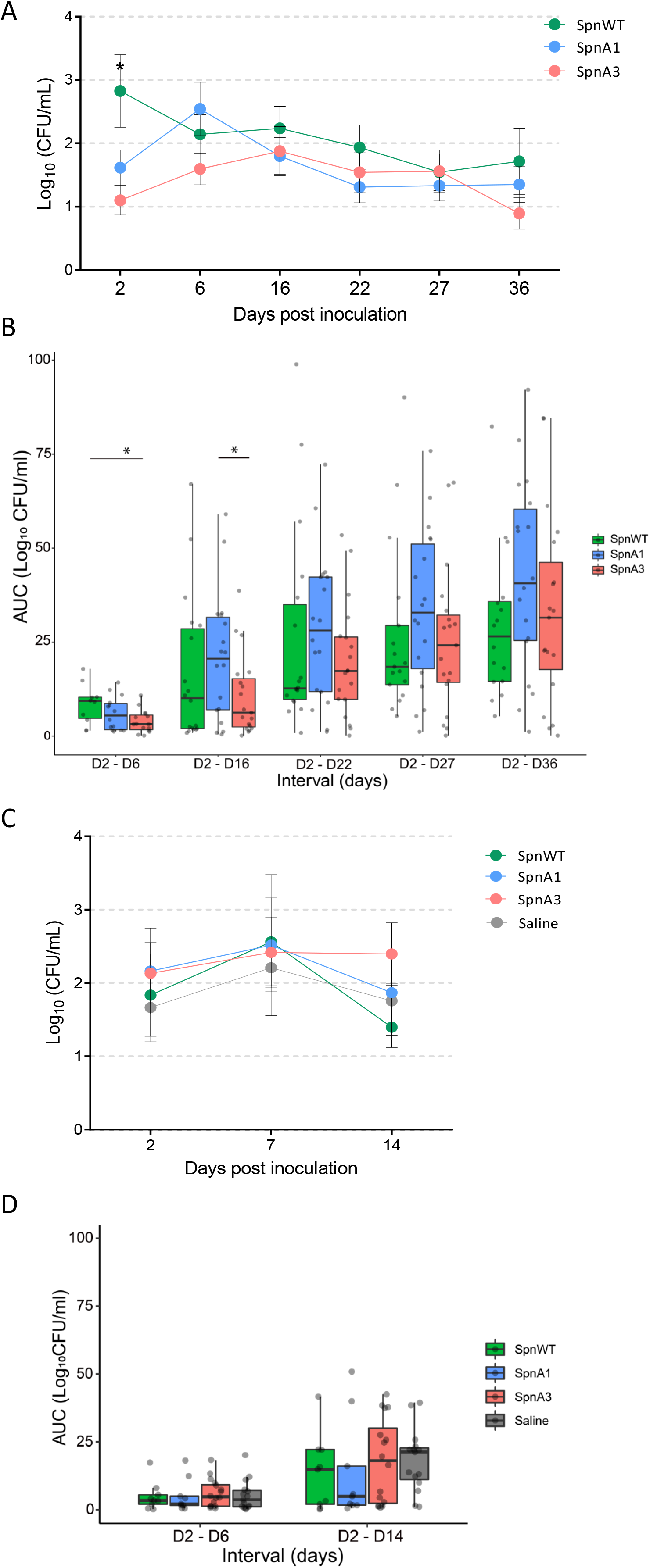
**(A-C)** Density dynamics after pneumococcal inoculation are calculated from classical microbiology [log10 (CFU/ml +1)]. Mean density of *Streptococcus pneumoniae* for each nasal wash time point amongst participants in whom SpnWT or attenuated strain was detectable at any time point at Stage I **(A)** or only SpnWT at any time point at Stage II (C). Bars represent SE of mean. **(B-D)** Area under the curve (AUC) of density-time intervals from D2 to D36 post inoculation with SpnWT or attenuated strains at Stage I **(B)** or D2 to D14 post challenge with SpnWT at Stage II (D). Box plot of median with interquartile range (IQR). * p <0.05, with one-way ANOVA test. Colour indicate the group comparison.

### Increased serum and mucosal antibody responses after colonisation with SpnWT, SpnA1 and SpnA3 mutant strains

Induction of antibodies after nasal inoculation was assessed in each group by whole cell ELISA using serum and nasal wash samples obtained on days -5, 2 (nasal wash only), 14 (serum only), 16 (nasal wash only) and 27 post first inoculation. Fold change to baseline is presented in Supplementary figure 3. At baseline (day -5) there were no differences in serum or nasal anti-Spn IgG levels among the SpnWT, SpnA1 and SpnA3 groups or among participants who were successfully colonised compared to those who were not colonised (Figure 3A). In non-colonised participants from all three Spn groups neither serum nor nasal anti-Spn IgG levels increased between baseline and day 27. In successfully colonised participants after the first and/or booster inoculation, serum anti-Spn IgG levels at day 27 were substantially increased compared to baseline in both SpnWT and SpnA1 groups (Figure 3A), with a trend towards an increase in those colonised with SpnA3.

**Figure 3.**
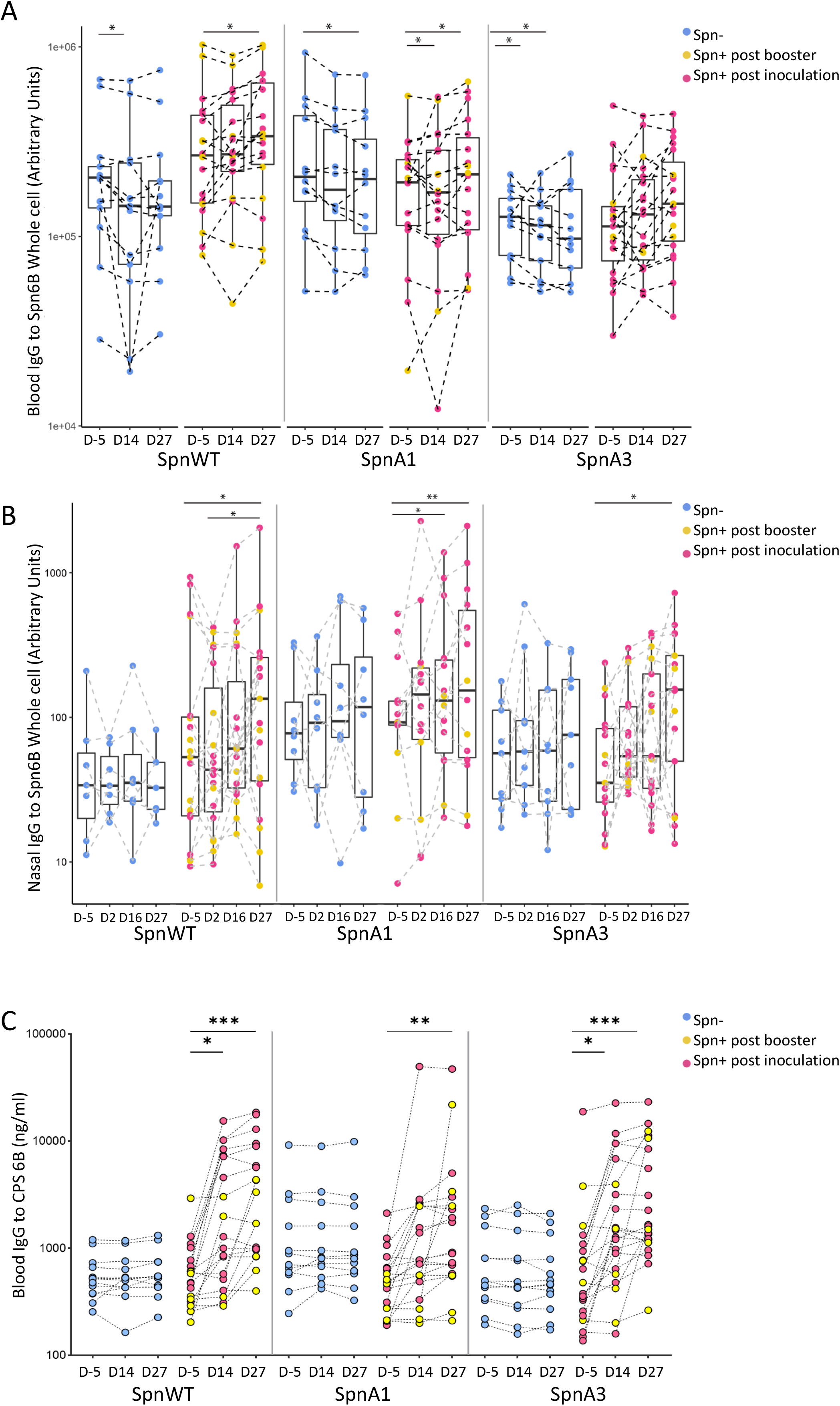
Anti-pneumococcal antibody induction after inoculation and booster dose with SpnWT or an attenuated strain. Kinetics of (**A**) serum and (**B**) nasal IgG levels to whole cell SpnWT and (**C**) serum IgG levels to SPS6B in study participants inoculated with SpnWT (n=31, 13 Spn- and 18 Spn+), A1 strain (n=30, 12 Spn- and 18 Spn+) or A3 (n=32, 13 Spn- and 19 Spn+). Boxplots depicting median and IQR. In blue depicted those particiapnts who did not become colonised (Spn-), in pink individuals who become colonised after initial inoculation (Spn+ post inoculation) and in yellow individuals who became colonised after booster inoculation (Spn+ post booster). * p <0.05 Friedman test and corrected for multiple comparisons with Dunn’s test.

Day 27 nasal anti-Spn IgG levels were also increased in Spn-colonised participants compared to day -5 for all groups. In addition, for the SpnWT and SpnA1 groups, levels of Spn specific nasal wash IgG were increased at day 27 compared to day 2 and at day 16 compared to baseline, respectively (Figure 3B). Overall, the whole cell ELISA data showed increase of nasal anti-Spn IgG levels for all Spn groups, but systemic responses were improved to a greater extent in response to SpnWT or SpnA1 strains compared to SpnA3.

We also assessed systemic IgG responses to CPS-6B after nasal inoculation using samples obtained on days -5, 14 and 27. Anti-CPS-6B levels did not increase from baseline in non-colonised individuals for any experimentally challenged group (Figure 3C). Conversely, in Spn colonised participants anti-CPS-6B IgG levels increased significantly at day 27 compared to baseline (all groups; p<0.01), and also between baseline and day 14 (p<0.05) for participants challenged with SpnWT or SpnA3 (Figure 3C). These data demonstrate enhanced systemic and nasal IgG anti-CPS6B response in all 3 groups following colonisation.

### Detailed characterisation of serum IgG responses to colonisation using an Spn antigen array

To assess systemic IgG responses to specific antigens, an Spn antigen array of 2,629 protein antigens (representing 2,454 genes) and 24 capsular polysaccharide antigens was probed with day -5 and +27 sera from the ten colonised subjects from each of the SpnWT, SpnA1 and SpnA3 groups showing the greatest increase in ELISA whole cell IgG titres. The combined data for all three groups demonstrated increases in the intensity of IgG responses to 197 protein antigens in total including the RrgA2 (pilus protein), variants of diverse core loci (“DCL”) PspA, PspC, ZmpA and ZmpB, and multiple non-DCL proteins such as the capsule biosynthesis proteins wzg and Cps4A, penicillin-binding protein 2b (Pbp2b), pneumococcal histidine triad protein E (PhtE) and beta galactosidase BgaA (Figure 4A and C). When the mean difference between each group was analysed individually, only participants from the SpnWT group showed significant increases in IgG responses to multiple protein antigens (Figure 4B). Of note, SpnWT colonised but not SpnA1 or SpnA3 colonised participants had improved IgG responses to PiaA, an antigen for which the corresponding gene has been deleted from strains SpnA1 and SpnA3 (Figure 4C). Similarly, the combined data showed increased IgG responses to the serotype 6B CPS and to a lesser degree to the related serotype 6A CPS (Figure 4D). Data for individual groups showed a significant increase in serum IgG to 6B CPS in the SpnWT group and a trend to an increase for the SpnA1 and SpnA3 groups (Figure 4E). Overall, these data show that established colonisation increased IgG responses to multiple protein antigens and the 6B capsule, with these responses being more pronounced in the SpnWT group. The differences in intensity of antibody responses per group may rely on the sera investigation from only ten Spn colonised participants per group.

**Figure 4.**
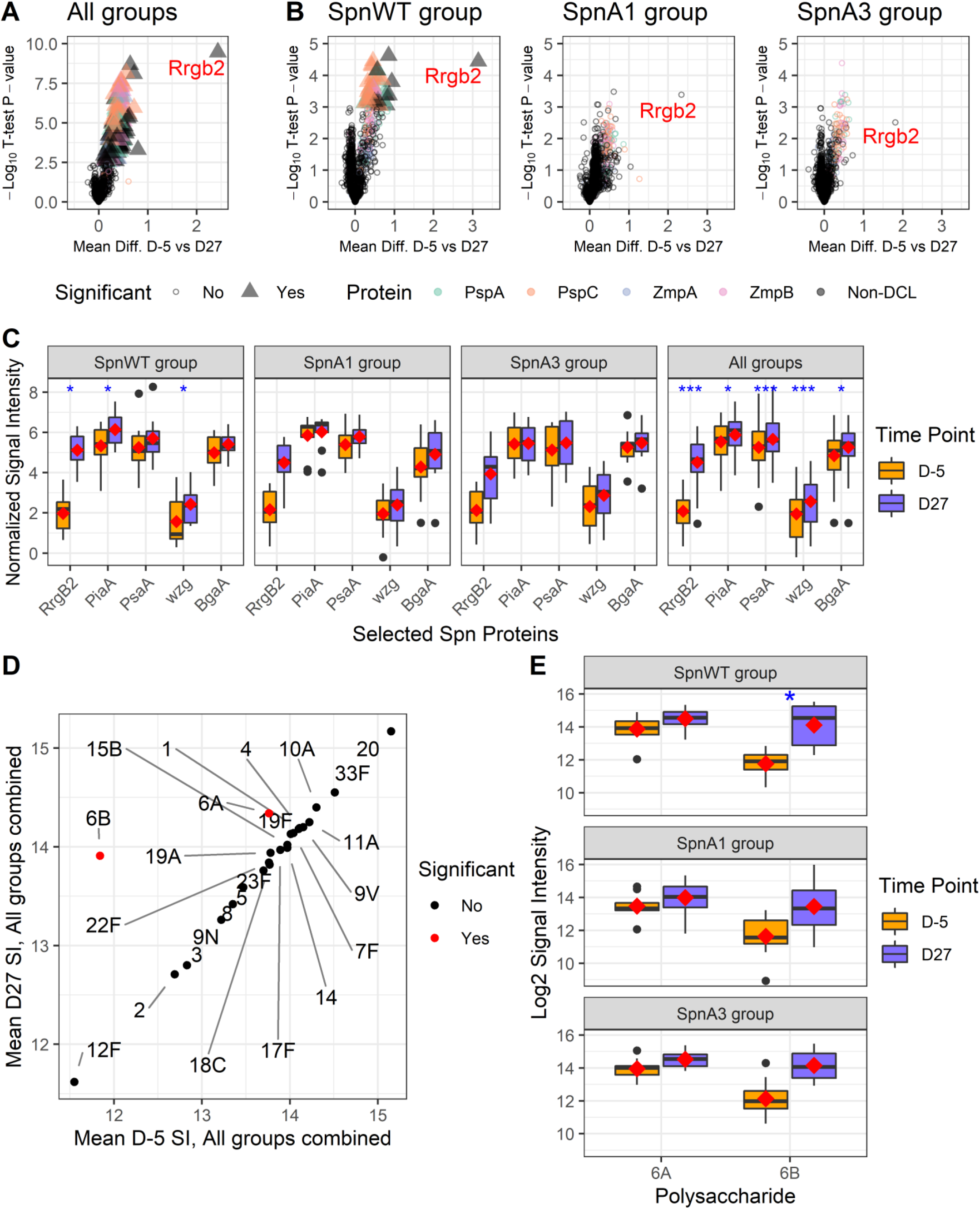
Proteome-wide and anti-capsular polysaccharide antibody induction after inoculation and booster dose with SpnWT or an attenuated strain. Responses against 2,629 pneumococcal proteins representing 2,454 unique genes are shown in volcano plots for ten colonised participants from each of the SpnWT, A1 and A3 groups showing the greatest increase in ELISA whole cell IgG (**A**) and for each of the groups individually (**B**). The mean difference between baseline (D-5) and post-inoculation (D27) are shown on the x-axis, and the inverse log10 t-test P-values are shown on the y-axis. Significantly different antigens are show in triangles, and points are colored if the protein was a variant of the diverse core loci (“DCL”) PspA, PspC, ZmpA or ZmpB. Boxplots show the distribution of signals for selected antigens with the mean overlaid as red diamonds (**C**). For capsular polysaccharides, the scatterplot (**D**) shows the range of responses pre- and post-inoculation with any Spn and significant increases in IgG only for serotypes 6B and 6A. (**E**) The boxplot of individual group responses to capsulare polysaccharides 6A and 6B. Blue asterisks represent p-values adjusted for the false discovery rate of <0.05 (*), <0.005 (**) and <0.0005 (***).

### SpnWT and SpnA1 challenge were associated with reduced colonisation rates when rechallenged with the WT strain

After rechallenge with the SpnWT strain at 6 months (range 129-174 days), the recolonisation rate for the SpnA3 (16/32, 50%) was similar to the saline group (15/32, 46.9%). In contrast, the recolonisation rates were lower for participants from the SpnWT (9/31, 29%) and SpnA1 (9/30, 30%) groups (Table 1). Compared to the saline group the relative risk of re-colonisation on days 2, 6, and 14 post-challenge with WT were lower for both the SpnWT and A1 groups but not the A3 group, with the difference achieving statistical significance on day 14 (A1 RR=0.33 [0.12-0.89, p=0.02], WT RR= 0.38 [0.16-0.95, p=0.03] (Table 1 and 2). For participants successfully recolonised with WT in Stage II densities of colonisation were comparable to those seen in Stage I (Figure 2C). Although AUCs for WT recolonisation of the SpnWT and A1 groups were generally reduced compared to the saline group, the variation between participants prevented these differences reaching statistical significance (Figure 2D and Supplementary Table 5 and 6). The duration of WT recolonisation was shorter for the SpnA1 (but not the WT) group (6 days [IQR 2-14]) compared to the saline group (14 days [IQR 14-14] (GEE model, p=0.03) (Supplementary Table 3).

**Table 2:** GEE model analysis of Stage II SpnWT re-colonisation rates for the Stage I WT, A1, and A3 groups. Data are expressed as a ratio of re-colonisation rates in the saline group; statistically significant differences are highlighted in bold

| Stage II | SpnA1 | SpnA3 | SpnWT |
| --- | --- | --- | --- |
| Day 2 | 0.60 (0.27,1.32), P=0.20 | 1.21(0.68,2.16), P=0.51 | 0.67(0.32,1.40), P=0.28 |
| Day 6 | 0.50(0.20,1.27), P=0.14 | 0.94(0.47,1.89), P=0.85 | 0.47(0.18,1.19), P=0.11 |
| Day 14 | **0.33(0.12,0.89), P=0.02** | 0.79(0.41,1.53), P=0.49 | **0.38(0.16,0.95), P=0.03** |
| any time point | 0.64(0.33,1.24), P=0.18 | 1.07(0.64,1.77), P=0.80 | 0.62(0.32,1.20), P=0.15 |

A post hoc analysis evaluated re-colonisation rates according to whether participants were colonised with Spn or not during Stage I (Figure 1B). Re-colonisation rates for Spn-colonised or non-colonised participants in Stage I were similar in SpnWT (6/18, 33% vs 3/13, 23%, p=0.41) and SpnA1 groups (5/18, 28% vs 4/12, 33%, p=0.52). However, for the SpnA3 group re-colonisation was more frequent in previously non-colonised participants than those previously successfully colonised (9/13, 69%, vs 7/19, 37%, p=0.07) (Figure 1B).

## Discussion

Controlled human infection models are useful platforms for testing of candidate vaccines and defining the host mechanisms of protection under controlled conditions using relatively small numbers of participants (32). Using an early phase clinical trial in the established EHPC model we have now assessed the efficacy of nasopharyngeal administration of genetically modified Spn strains attenuated in virulence at protecting against re-colonisation with SpnWT. The results confirmed that human colonisation with genetically modified low virulence S. pneumoniae strains (24) can be achieved without adverse effects, and can improve humoral immunity to pneumococcus. Importantly, administration of the mutant SpnA1 reduced the rates of re-colonisation 6 months later from 47% to 30%, an almost identical result to prior administration of the WT strain. Previous data suggested that prior colonisation with SpnWT prevented re-colonisation in 100% of participants (22), whereas in this trial the efficacy was significantly lower at 70%. However, as colonisation is the pre-requisite for invasive infection (18, 21, 22, 24, 37), if this level of protection against colonisation after nasal administration of strain SpnA1 was serotype independent, this could reduce the incidence of pneumonia by approximately a third. This is a higher efficacy that achieved by vaccination of adults with either the 13-valent pneumococcal conjugated vaccine (8) or the unconjugated pneumococcal vaccine (38). Administration of the mutant SpnA1 could also result in additional protection through its effect on improving protective IgG responses to Spn, and potentially other effects on immunity not assessed in this study including improved T cell responses and alveolar macrophage function (14, 15, 22, 23, 32, 39). Recent data on the efficacy of nasal administration of *N. lactamica* have shown similar protective effects against *N. meningitidis*, with induction of B cell responses and an approximately 56% decrease in *N. meningitidis* nasopharyngeal colonisation affecting multiple strain genotypes (25, 26). Future experiments will investigate whether nasal administration of attenuated *Spn* strains such as SpnA1 can prevent colonisation with heterologous *Spn* strains and in participants more susceptible to *Spn* infection due to age or underlying comorbidities. Furthermore, the attenuated virulence of SpnA1 means a higher dose inoculum can be given which may induce stronger protection against subsequent Spn infection.

An important observation from this study is that the colonisation phenotypes of the mutant strains in mouse models were replicated in the EHPC model; strains A1 and A3 are able to colonise both mouse and human nasopharynx, whereas the unencapsulated strain A2 which has an impaired ability to colonise mice (24) also failed to achieve high colonisation rates in humans. This is the first study to use deliberate infection of humans with genetically modified *Spn*, and the results provide confidence that specific *Spn* mutant phenotypes identified using mouse models are relevant during human infection. Future experiments using the EHPC model and genetically manipulated *Spn* strains could be highly informative about key host-bacterial interactions during nasopahryngeal colonisation.

Despite previous intense study, the actual mechanism(s) by which prior administration of Spn prevents re-colonisation in human infection remains unclear (23, 28, 29). Candidate mechanisms include local antibody and/or T cell cellular responses to protein antigens, improved local innate immunity, both of which would provide serotype independent protection, or antibody to capsular antigens that would provide serotype specific protection. We observed increased systemic anti-CPS-6B specific responses in colonised participants for all three groups, and increased mucosal and systemic levels of whole cell ELISA IgG in participants colonised with SpnWT, SpnA1, and to a lesser extent SpnA3. Pooled data obtained using an *Spn* pangenome protein array demonstrated colonisation was associated with increased serum IgG responses to nearly 200 proteins, confirming colonisation has a significant immunostimulatory effect. The pattern of antibody response to Spn protein antigens vary markedly between individuals (34), and our pangenome antibody data were too underpowered to interpret the results obtained separately for the SpnWT, SpnA1 and SpnA3 groups. In addition, T cells and other cellular responses were not analysed in this study. These will be important areas of future study, especially as in the SpnWT and SpnA1 groups improved antibody responses were limited to successfully colonised individuals, there was equal levels of protection against recolonisation between uncolonised and colonised, participants. Although the mouse data showed previous colonisation with either strain A1 and or A3 was protective (24), after human challenge only SpnA1 was protective. In mouse models a longer duration of *Spn* nasopharyngeal colonisation increased immune responses (11). Hence the lower density of colonisation SpnA3 during the first few days after the first nasal administration compared to SpnWT and SpnA1 could have adversely affected immune responses to this strain. The failure of the SpnA3 strain to prevent recolonisation provides a useful tool to help identify the mechanism(s) preventing recolonisation. A detailed analysis using the EHPC model of the differences in epithelial and immune responses between strains SpnA1 and SpnA3 should identify potential correlates for prevention of re-colonisation, and which in turn could indicate how further genetic manipulation of live attenuated Spn mutant strains could enhance their protective effects.

## Conclusion

To conclude, using a human challenge model we have shown that nasal administration of the live attenuated genetically modified Spn strain A1 partially protects against Spn recolonisation and improves systemic and nasal antibody responses. Our data demonstrate nasal administration of genetically modified S. pneumoniae can be a tool to enhance immunity to pneumococcus, and which could contribute to future strategies for preventing Spn infections.

## Data Availability

All data produced in the present study are available upon reasonable request to the authors

## Acknowledgments

We would like to thank the Data Monitoring and Safety Committee (Prof R Read – chair (University of Southampton), Dr M Gibani (University of Oxford), Prof B Farragher (Liverpool School of Tropical Medicine). We would also like to acknowledge support of the Respiratory department at the Royal Liverpool Hospital (M Gautam, J Hadcroft, P Deegan, L Chishimba, G Jones, S Zaidi, C Smyth, W Kent), the wider LSTM team, NIHR CRN NWC, our research ambassadors and all of the trial participants.

## Notes

### Competing Interest Statement

The authors have declared no competing interest.

### Clinical Trial

ISRCTN22467293

### Funding Statement

This study was funded by MRC

