## Supplementary material for "A Randomised Controlled Trial of Nasal Immunisation with Live Virulence Attenuated *Streptococcus pneumoniae* Strains using Human Infection Challenge"

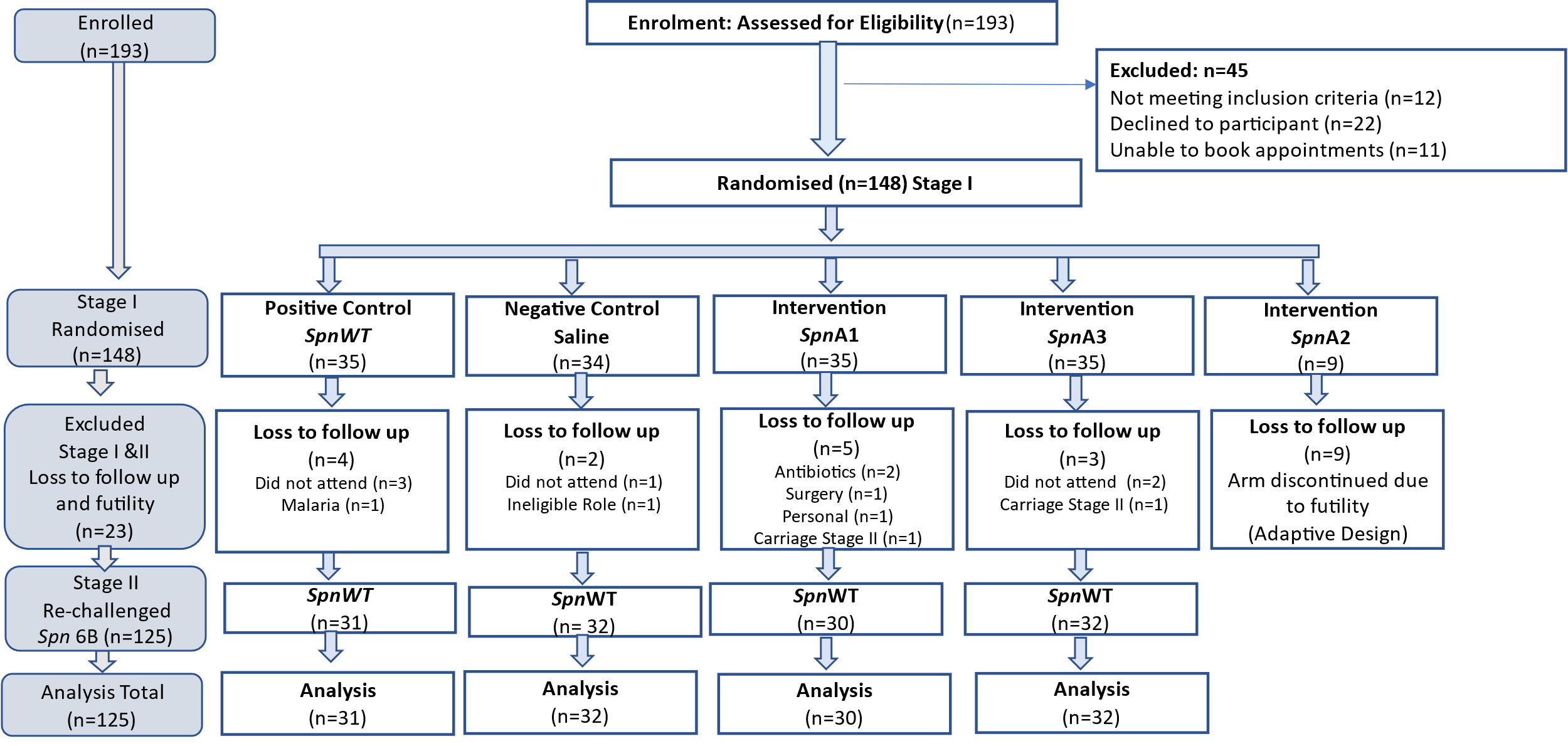

**Supplementary figure 1.** Consort flow chart

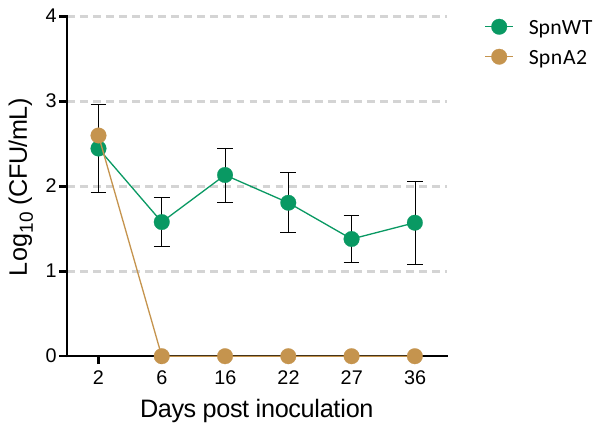

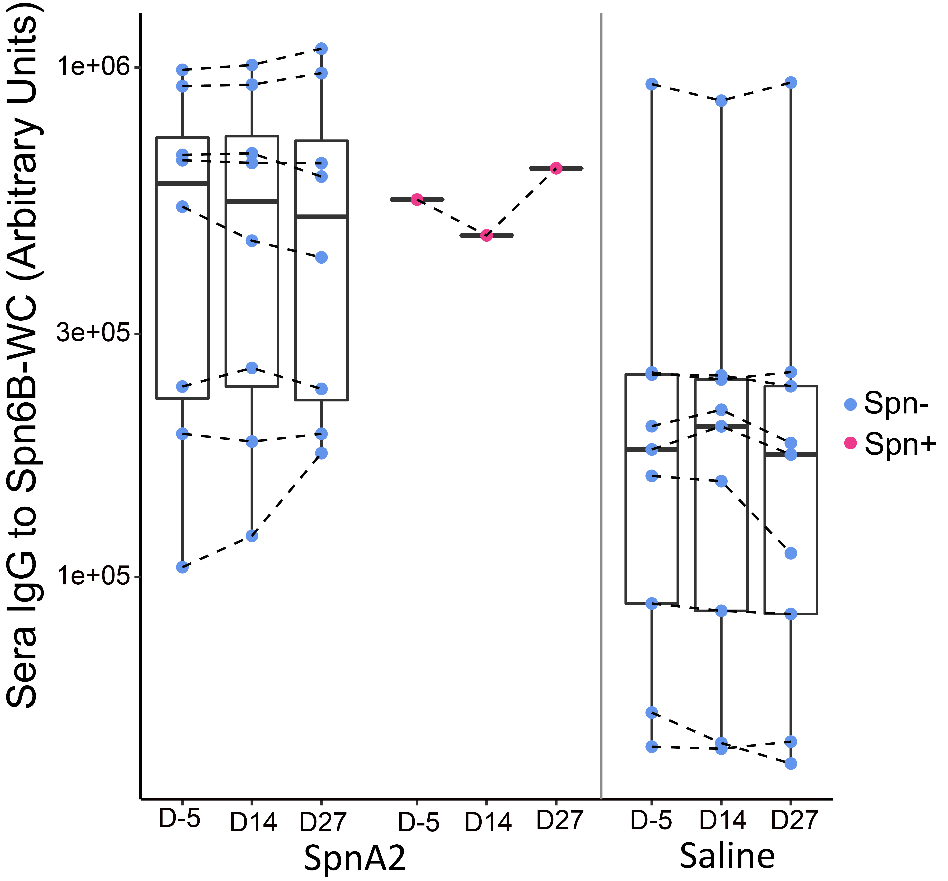

**Supplementary figure 2.** Kinetics of serum IgG levels to whole cell of Spn6B wild type measured in study participants inoculated with attenuated strain 2 (A2) (n=9) divided in pneumococcal colonised (n=8) and non-colonised (n=1) and in a subset of participants (n=8) inoculated with saline (control group). Inset: Density of the 1 individual colonised with A2, detected at only D2 post inoculation alongside with mean densities of Spn6B wild type throughout phase I time points.

A

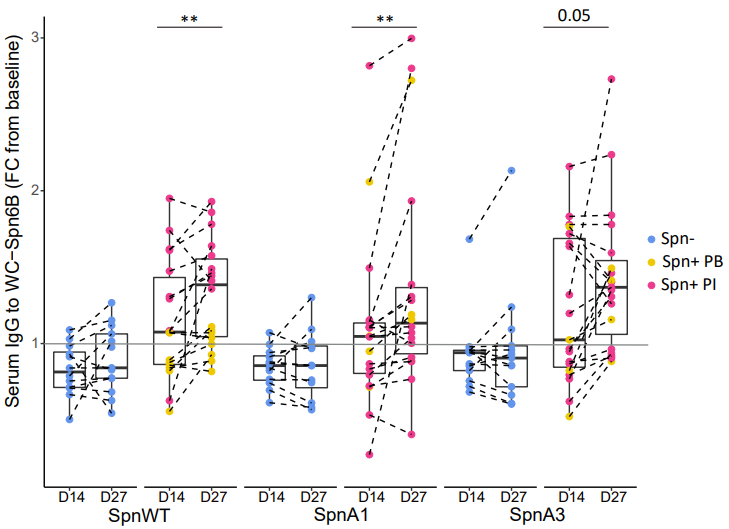

B

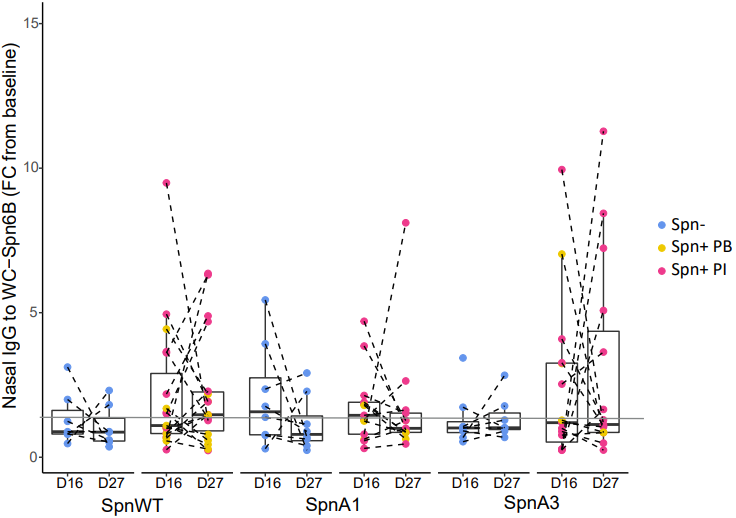

**Supplementary figure 3.** Fold change of IgG to whole cell of Spn6B at D14 and D27 from baseline in sera (A) and at D16 and D27 from baseline in nasal wash (B). Wilcoxon test was used for the comparison of antibody FC within the same group *p≤0.05. Kruskal-wallis test was used for the comparison of IgG FC at D14/D16 or D27 between the 3 different groups. No stat sign difference within the same group or amongst groups.

**Supplementary Tables**

**Supplementary Table 1:** Participants excluded due to having natural *S. pneumoniae* colonization of the nasopharynx.

| **Participant Number** | | | **Dose** **CFU/ul** | | | | **pre-inoculation** | | | | | | **d2** | | **d6** | | | **Booster CFU/ul** | | | | **d16** | | | **d22** | | | **d27** | **d36** |
| --- | --- | --- | --- | --- | --- | --- | --- | --- | --- | --- | --- | --- | --- | --- | --- | --- | --- | --- | --- | --- | --- | --- | --- | --- | --- | --- | --- | --- | --- |
| Volunteer 1 | | | 70000 | | | | Spn9 | | | | | | ND | | Spn9 | | | 65167 | | | | Spn9 | | | ND | | | Spn9 | ND |
| Volunteer 2 | | | 80333 | | | | ND | | | | | | ND | | ND | | | 78167 | | | | ND | | | ND | | | ND | ND |
| Volunteer 3 | | | 75167 | | | | Spn19 | | | | | | Spn19 | | Spn19 | | | 76167 | | | | ND | | | Spn19 | | | Spn19 | N/A |
| Volunteer 4 | | | 78834 | | | | Spn33 | | | | | | Spn33 | | Spn33 | | | 88500 | | | | Spn33 | | | ND | | | ND | Spn33 |
| Volunteer 5 | | | NA | | | | ND | | | | | | ND | | ND | | | 0 | | | | ND | | | ND | | | ND | Spn3 |
| Volunteer 6 | | | NA | | | | Spn10 | | | | | | ND | | ND | | | 0 | | | | ND | | | ND | | | ND | ND |
| Volunteer 7 | | | NA | | | | ND | | | | | | ND | | ND | | | 0 | | | | ND | | | ND | | | ND | ND |
| Volunteer 8 | | | 75166 | | | | Spn8 | | | | | | ND | | ND | | | 82500 | | | | ND | | | ND | | | ND | ND |
| Volunteer 9 | | | 89833 | | | | ND | | | | | | ND | | NVT | | | 91166 | | | | ND | | | ND | | | ND | ND |
| **Participant Number** | | | | **Group** | | | | **dose CFU/ul)** | | | **pre-challenge** | | | | | | **d2** | | **d6** | | | | **d14** | | | |  |  |  |
| Volunteer 10 | | | | 6BWT | | | | 78667 | | | Spn11 | | | | | | ND | | ND | | | | ND | | | |  |  |  |
| Volunteer 11 | | | | 6BWT | | | | 81167 | | | ND | | | | | | ND | | ND | | | | ND | | | |  |  |  |
| Volunteer 12 | | | | 6BWT | | | | 78500 | | | ND | | | | | | ND | | ND | | | | ND | | | |  |  |  |
| Volunteer 13 | | | | 6BWT | | | | 77667 | | | ND | | | | | | ND | | ND | | | | ND | | | |  |  |  |
| Volunteer 14 | | | | 6BWT | | | | 79833 | | | NVT | | | | | | ND | | ND | | | | ND | | | |  |  |  |
| Volunteer 15 | | | | 6BWT | | | | 84334 | | | ND | | | | | | ND | | ND | | | | ND | | | |  |  |  |
| Volunteer 16 | | | | 6BWT | | | | 78167 | | | ND | | | | | | ND | | ND | | | | ND | | | |  |  |  |
| Volunteer 17 | | | | 6BWT | | | | 86334 | | | ND | | | | | | ND | | ND | | | | ND | | | |  |  |  |
| Volunteer 18 | | | | 6BWT | | | | 84667 | | | Spn11 | | | | | | Spn11 | | ND | | | | ND | | | |  |  |  |

**Supplementary Table 2**: Demographics of the participants and experimental details

|  |  | SpnWT  **(N=31)** | SpnA1  **(N=30)** | SpnA3  **(N=32)** | Saline  **(N=32)** |
| --- | --- | --- | --- | --- | --- |
| Median age (IQR) – yr | | 22.0  (20.0-23.0) | 21.0  (19.0-27.0) | 21.5  (19.0-24.5) | 22.0  (20.5-25.0) |
| Female – no. (%) | | 21 (67.7%) | 15 (50%) | 23 (71.9%) | 21 (65.6%) |
| Median dose (IQR) – CFU/nostril | | 79667  (75500-81167) | 84333  (81666-87166) | 73333  (71667-76166) | 0  (0-0) |
| Median dose booster (IQR) – CFU/nostril | | 78167  (74667-80499) | 86000  (82499-88000) | 76834  (74834-79167) | 0  (0-0) |
| Median dose Stage II (IQR) – CFU/nostril | | 80833  (77917-83667) | 79750  (77167-83833) | 82583  (78500-85667) | 80083  (78167-83667) |
| Time to Booster (days) Mean SD | | 14.0  (14.0-14.0) | 14.0  (14.00-14.0) | 14.0  (14.00-14.0) | 14.0  (14.00-14.0) |
| Time to Stage II inoculation (days) Mean SD | | 154.0  (140.0-168.0) | 157.5  (148.0-174.0) | 146.0  (129.0-160.5) | 157.0  (140.0-168.5) |

**Supplementary Table 3**: Colonisation duration (Median days, IQR) in Stage I and Stage II

|  | SpnWT  **(N=31)** | SpnA1  **(N=30)** | SpnA3  **(N=32)** | Saline  **(N=32)** | |
| --- | --- | --- | --- | --- | --- |
| Stage I | N=18 | N=18 | N=19 | | - |
|  | 36 (22-36) | 36 (27-36) | 36 (27-36) | | - |
| Stage II | N=9 | N=9 | N=16 | | N=15 |
|  | 14 (2-14) | 6 (2-14) | 14 (2-14) | | 14 (14-14) |
| GEE compared to Saline | P=0.09 | P=0.03 | P=0.16 | | - |

**Supplementary Table 4**: *S. pneumoniae* colonisation density (log_10_ CFU/ml) stratified by time point

|  |  | | SpnWT  N=31 | | SpnA1  N=30 | | | SpnA3  N=32 | | | Saline  N=32 | | |
| --- | --- | --- | --- | --- | --- | --- | --- | --- | --- | --- | --- | --- | --- |
| **Stage I** | | **n** | | **Mean ± SD** | | **n** | **Mean ± SD** | | **n** | **Mean ± SD** | | **n** | **Mean ± SD** |
| d2 | | 10 | | 2.45 ± 1.64 | | 9 | 1.53 ± 0.90 | | 13 | 1.08 ± 0.78 | | - | - |
| d6 | | 13 | | 1.82 ± 1.05 | | 14 | 2.42 ± 1.61 | | 12 | 1.53 ± 0.82 | | - | - |
| d16 | | 19 | | 2.13 ± 1.40 | | 17 | 1.80 ± 1.20 | | 13 | 1.89 ± 1.29 | | - | - |
| d22 | | 14 | | 1.81 ± 1.32 | | 16 | 1.31 ± 1.00 | | 15 | 1.51 ± 1.12 | | - | - |
| d27 | | 14 | | 1.38 ± 1.03 | | 14 | 1.33 ± 0.91 | | 14 | 1.51 ± 1.18 | | - | - |
| d36 | | 11 | | 1.57 ± 1.63 | | 12 | 1.45 ± 0.99 | | 13 | 0.92 ± 0.83 | | - | - |
| **Stage II** | |  | |  | |  |  | |  |  | |  |  |
| d2 | | 8 | | 1.83 ± 1.59 | | 7 | 2.38 ± 1.67 | | 15 | 2.13 ± 1.63 | | 12 | 1.66 ± 1.62 |
| d6 | | 5 | | 2.45 ± 1.34 | | 5 | 2.51 ± 2.15 | | 10 | 2.42 ± 1.53 | | 11 | 2.21 ± 1.08 |
| d14 | | 5 | | 1.55 ± 0.64 | | 4 | 1.87 ± 1.16 | | 10 | 2.39 ± 1.35 | | 13 | 1.76 ± 0.85 |

**Supplementary Table 5:** Area under the density curve for each interval *

|  | SpnWT | | SpnA1 | | SpnA3 | | Saline | |
| --- | --- | --- | --- | --- | --- | --- | --- | --- |
|  | **n** | **Mean ± SD** | **n** | **Mean ± SD** | **n** | **Mean ± SD** | **n** | **Mean ± SD** |
| **Stage I** |  |  |  |  |  |  |  |  |
| d2 – d6 | 13 | 7.13 ± 5.39 | 14 | 6.02 ± 4.48 | 15 | 3.91 ± 3.02 | - | - |
| d2 – d16 | 21 | 15.68 ± 18.37 | 17 | 22.05 ± 16.77 | 19 | 10.59 ± 10.78 | - | - |
| d2 – d22 | 21 | 23.91 ± 25.50 | 18 | 28.78 ± 20.04 | 19 | 19.84 ± 15.06 | - | - |
| d2 - d27 | 21 | 29.46 ± 29.41 | 18 | 33.95 ± 21.22 | 19 | 25.56 ± 19.30 | - | - |
| d2 – d36 | 21 | 37.59 ± 36.21 | 18 | 42.51± 25.94 | 19 | 33.14 ± 25.36 | - | - |
| **Stage II** |  |  |  |  |  |  |  |  |
| d2 – d6 | 9 | 4.96 ± 5.25 | 9 | 5.18 ± 6.09 | 16 | 6.13 ± 5.15 | 15 | 5.35 ± 5.55 |
| d2 - d14 | 9 | 13.65 ± 13.83 | 9 | 13.76 ± 18.74 | 16 | 18.07 ± 16.63 | 15 | 18.24 ± 11.48 |

* Mean ± SD area under the log10+1-transformed density curve (AUC) was calculated for participants positive for Spn based on classical microbiology or molecular methods at any time point. n = the number of positive volunteers for whom an AUC could be calculated over that interval using the trapezoid rule. Missing density values for time points flanked by known density values were interpolated with the mean of these known values. Sequential missing values and/or missing values at the end of a given interval were extrapolated using the average change over that interval stratified by vaccine.

**Supplementary Table 6**: GEE analysis Area under the density curve

|  | SpnA1 | SpnA3 | SpnWT |
| --- | --- | --- | --- |
| d2-6 | -0.17(-4.50,4.16) P=0.93 | 0.78 (-2.91,4.48) P=0.67 | -0.39 (-4.72,3.95) P=0.86 |
| d2-14 | -4.48(-16.19,7.23) P=0.45 | -0.17(-10.15,9.82) P=0.97 | -4.59(-16.30,7.13) P=0.44 |

**Supplementary methods:**

Inclusion and Exclusion criteria for the study

**Inclusion**

- Healthy adult
- Age 18 – 50 years
- Capacity to give informed consent
- Ability to speak fluent English

**Exclusion**:**

- **Research participant:**
  - Currently involved in another study unless observational or non-interventional except for the EHPC bronchoscopy study*
  - Participant in a previous EHPC trial (that result in nasal inoculation or carriage)
- **Vaccination:** pneumococcal vaccination (routine in UK babies born since 2005 or US 2001)
- **Allergic:** to penicillin, amoxicillin
- **Health history:**
  - Chronic ill health including, immunosuppressive history, diabetes, asthma (on regular medication), recurrent otitis media or other respiratory disease
  - Medication that may affect the immune system or clotting e.g. steroids, inflammation altering (e.g. nasal steroids, roacutane or aspirin)
  - Recent antibiotics (within the last 4 weeks or long term for known active chronic infection)
  - Splenectomy
  - Current acute severe febrile illness
  - Major pneumococcal illness requiring hospitalization
  - Other conditions considered by the clinical team as a concern for participant safety or integrity of the study
- **Direct caring role or close contact** with individuals at higher risk of infection
  - Children under 5 years age
  - Chronic ill health or immunosuppressed adults
  - Adults over the age of 75 years
- **Smoker:**
  - Current or ex-smoker (regular cigarettes, e-cigarette/vaping and recreational drugs) in the last 6 months
  - Previous significant smoking history - more than 20 cigarettes per day for 10 years or the equivalent (>10 pack years)
- **Women of child-bearing potential (WOCBP**) who are:
  - not deemed to have sufficient /effective birth control or confirmed abstinence
  - pregnant
  - **History of drug or alcohol abuse** (at discretion of the clinician)
  - **Overseas travel planned** in follow up period of Stage I or Stage II
